# Systematic Data Fitness Assessment Improves Validity and Replicability of Research Using Real-World Data

**DOI:** 10.64898/2026.08.05.26359818

**Authors:** Hanieh Razzaghi, Kaleigh Wieand, Annabel Pinkney, Charles Bailey

## Abstract

Research replication in real-world data is essential to build trust in evidence from clinical studies. However, methods for conducting and reporting these efforts are lacking, particularly related to data fitness and limitations. We demonstrate the importance of incorporating systematic fitness testing by replicating a single-center observational study of hydroxyurea for children with severe sickle cell disease (SS/Sβ0) in a multi-institutional learning network using EHR data (PEDSnet). An AS-IS arm applied the original study’s criteria with no major data quality adjustments, while a Data Fitness Enhanced (DFE) arm used systematic data fitness assessment to inform adjustments to cohort eligibility criteria and variable definitions; both arms then replicated the original study’s primary analyses. Data quality checks in the DFE arm refined cohort accuracy and improved hydroxyurea capture, drug era computation, and hematology specialist mapping. The DFE cohort produced average treatment effects with higher face validity and greater concordance with the original study (e.g., change in ED visits: -0.44 (CI -0.60, -0.26) versus -0.36 (CI -0.57, -0.16) in the original study) than the AS-IS cohort (-0.08 (CI -0.26, 0.09)), which yielded several implausible results. These findings show that superficially plausible cohort characteristics do not guarantee valid results without transparent, systematic data fitness assessment.

## INTRODUCTION

Secondary use of clinical data offers unprecedented opportunities to conduct large-scale research and derive insights into patient care. These studies are more accessible and less costly than clinical trials, national surveys, or longitudinal cohort collection. However, clinical data are heterogeneous and complex, requiring multidisciplinary teams to interpret them rigorously and reduce bias in research. Multi-institutional clinical research networks have attempted to address this challenge by implementing comprehensive data quality (DQ) programs^1–7^. While useful for network governance and maintaining general usability standards, these programs are insufficient for study-specific analyses where complex clinical cohorts and variables exceed the scope of routine network DQ assessment^8, 9^.

Study-specific data fitness assessments are often conducted ad hoc, and their results can introduce significant and unforeseen delays in analysis^10–12^. DQ findings may occasionally render results unusable, but more commonly require researchers to alter study design or variable definitions. These changes do not compromise scientific integrity, but without transparency and standardized reporting approaches, they can limit generalizability and reproducibility. While peer-reviewed journals require transparency in statistical methods, they do not mandate reporting of data quality issues or the steps investigators took to curate the final study dataset. These are often critical ingredients in not just understanding the original study, but for replicating work in future studies.

To address both the need for comparable approaches and the resource cost of study-specific testing, our research group has recently described a set of standardized modules that can be rapidly applied to assess data fitness for an intended analysis^9^. The purpose of this study is to demonstrate the use of standardized study-specific data fitness assessments for both robust evidence generation and to support research replication and generalizability. We replicate a study conducted at Children’s Healthcare of Atlanta (CHOA) examining the long-term effectiveness of hydroxyurea in treating pediatric patients with more severe forms of sickle cell disease^13^. We selected this study because it addresses an important area of investigation — real-world drug effectiveness — in a patient population that is heterogeneous and a disease whose severity is difficult to predict^14^. Notably, while CHOA used electronic health record (EHR) data as the foundation for their analyses, the authors supplemented these data with an institution-specific patient registry and chart reviews, described in more detail below. This distinction underscores the challenge of replicating such studies in multi-institutional databases, where these supplementary resources are typically unavailable or prohibitively expensive. Finally, the original investigators published their analytic code on GitHub with thorough documentation, further enabling replication across institutions and databases.

Our objective was to demonstrate the impact of standardized, transparent data fitness assessment on the replicability of real-world evidence generated from multi-institutional EHR data. Using PEDSnet, we replicated a single-center study of hydroxyurea’s long-term effectiveness in children with severe sickle cell disease under two conditions: a replication that applied the original study’s inclusion criteria and variable definitions without major modification, and a replication informed by systematic, standardized data fitness assessment that permitted adjustments to cohort and variable definitions based on the characteristics of the underlying data resource. We hypothesized that the data fitness-informed replication would produce results with greater face validity and clinical outcomes, demonstrating that rigorous, transparent data fitness assessment is necessary to generate valid, replicable evidence from secondary use of EHR data.

## METHODS

### Description of the original Study

The original research study published in July 2025 comprised patients under the age of 18 with more severe variants of Sickle Cell Disease (SCD), that is, those with SS/Sβ0 genotype. Patients eligible for the study were validated through a hospital-specific registry of verified patients with SS/Sβ0 and hemoglobin typing result validation. Children were excluded from analysis if they had < 3 total clinic visits and were censored at their last clinical visit if they went > 2 years without a clinical visit, underwent bone marrow transplant or gene therapy, or started chronic transfusion therapy or other disease modifying medication.

The primary exposure in the study was hydroxyurea use during a specific age-year, which was verified through manual review of clinical notes and prescription details from each office visit in the hematology department. Patients were classified as “taking” hydroxyurea if there was an active prescription at the visit, including newly issued or sufficient refill prescriptions to cover the visit time frame. Patients must have been covered by hydroxyurea for at least 50% of their visits in that year to be classified as exposed in the study.

The primary outcome was clinical utilization as measured by ED visits, hospitalization days per year, and annual average hemoglobin concentration. Baseline values were established by retrieving data from the initiation date of hydroxyurea treatment, with lab results ascertained to the closest clinic visit within one year prior to starting hydroxyurea. The authors conducted a difference-in-difference and dynamic event analysis for a cohort of 2,147 patients from 2010-2021. We focused on replicating the dynamic event analysis, which examined yearly outcomes to assess how hydroxyurea’s effects varied over time. All analyses were completed using the DiD package^15^.

### Study Sample

We used data from PEDSnet, the largest pediatric multi-institutional learning network in the US, to conduct our replication study^16, 17^. Data was drawn from the April 2025 [v57] version of the PEDSnet database and comprises 10 institutions. Our data extraction included anyone with a diagnosis code for SCD (all variants) and all their associated clinical data. Because no patient identifiers were present, this study was determined to be not human subjects research by the Institutional Review Board of the Children’s Hospital of Philadelphia.

### Study Design

We conducted the study using an *AS-IS* and a *Data Fitness Enhanced* (*DFE*) arm. For the *former*, we performed the analysis on our multi-site data, imposing the same inclusion and exclusion criteria as the original study with minor adaptations. For the *latter*, we performed data fitness assessments using our previously developed SQUBA data quality package^18^. The package contains a set of modules that can be configured for use across any dataset conformed to the OMOP or PCORnet^®^ common data models. These modules were developed to assess cohort identification, dataset fitness, and variable and codeset accuracy. We used a standardized and open-source DQ package to employ methods that are reproducible and transparent. For each round of analysis, we recorded the modules we implemented, lessons learned, action taken/next steps, implications for study design, and significance. Remediation of issues, variable definition changes, and other modifications were also recorded.

Importantly, in the DFE arm, we completed all our data quality work, which resulted in variable definition and study design changes described below in Results, prior to conducting the primary difference-in-difference dynamic event analysis. Similarly, the AS-IS arm was constructed without knowledge of the data quality outcome or study analysis results. These steps were taken to prevent bias based on known results from the AS-IS or interim DFE output.

Similar to the original study, we completed the primary analysis on utilization outcomes, including ED visits and hospitalization days, as well as average hemoglobin values. We built on the study team’s code, accessed through their well-documented GitHub repository^13^, using the same statistical packages as the original study.

### Data Quality Testing Process

We performed eight rounds of DQ checks, using combinations of eight SQUBA modules, which produced four successive patient cohorts, as shown in Figure 1. Each round focused on different aspects of DQ or was implemented based on an anomaly in the previous round.

**Figure 1.**
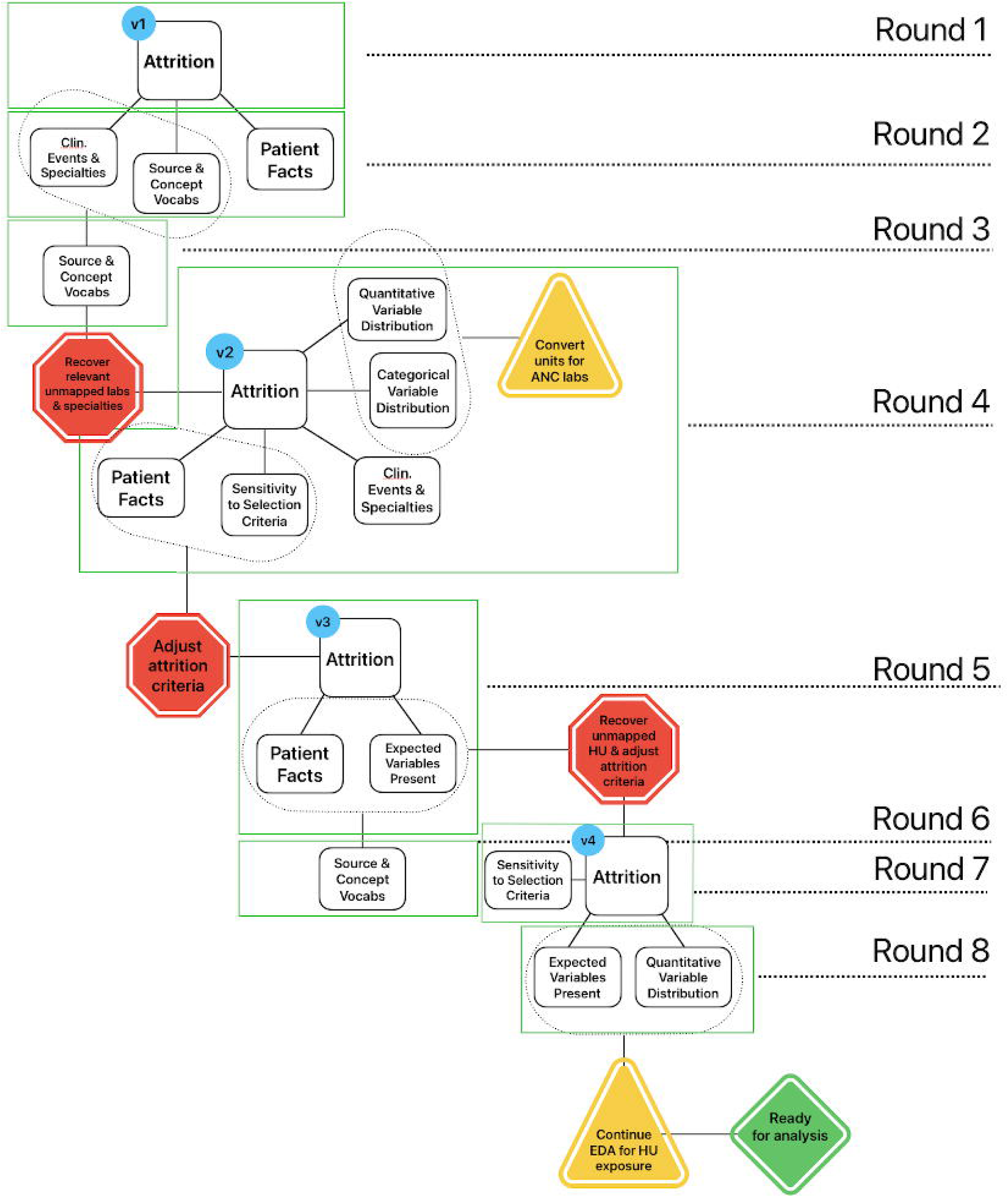
DQ Testing Process: Flow diagram of data quality testing process, including eight rounds across four different cohort revisions. Cohorts are denoted by v1, v2, v3, v4 and all begin with DQ evaluation of attrition. SQUBA module names used for DQ evaluation are in rectangles while adjustments and DQ remediation steps are in triangles and octagons.

Table 1 summarizes use of the DQ modules in the DFE arm. We prioritized review of study variables that were significant to the original research, such as confirmation of SS/Sβ^0^, evidence of hydroxyurea usage over time, and active engagement with a hematologist. The results of each round of DQ output informed the next modules to implement. For example, where we found poor capture of hematology specialty (Module: Clinical Events & Specialties), we investigated whether the information was recoverable in source data information that institutions submit (Module: Source & Concept Vocabularies). The goal of these iterations were to either make improvements to the data where possible or alter the study implementation based on proxy definitions or other methods to preserve the intent of the original study. For example, large scale multi-institutional data resources rarely have access to registries confirming SCD patients and hemoglobin typing result data may not be available at the institution where patients follow up with hematology care. As a result, we adjusted the study cohort definition for continuity of care and multiple instances of SCD-related encounters.

**Table 1.**

| **DQ Module** | **DQ Round(s)** | **Module Output** |
| --- | --- | --- |
| Attrition | 1,4,5,7 | • Individual attrition steps to detect potential data anomalies across institutions such as sudden drop offs<br>• Primary mechanism to identify downstream DQ issues and finalize study cohort |
| Clinical Events & Specialties | 2,4 | • Proportion of events that occur in visits with no provider specialty to evaluate missingness of specialty data<br>• Hematology specialty visit utilization to interrogate assumption that clinical events such as hydroxyurea prescription only occurred at mapped specialist visits |
| Clinical Fact Documentation | 2,4,5 | • Utilization of clinical event classes per person-year by visit type – e.g., outpatient procedures vs inpatient procedures – to highlight any major gaps across institutions<br>• Measures must be relevant to the study, such as frequency of hydroxyurea prescriptions and hemoglobin labs |
| Source & Concept Vocabularies | 2, 3, 7 | • Uncoded values for critical study variables from source systems resulting in missingness of data, such as specialty or SCD diagnostic lab testing |
| Quantitative Variable Distribution | 4,8 | • Distribution of various lab values to understand heterogeneity of results and quality across participating institutions<br>• Distribution of drug metadata to determine how to account for drug prescription continuity and gaps |
| Categorical Variable Distribution | 4 | • Distribution of units for MCV, ANC, and hemoglobin to aid with result interpretation |
| Sensitivity to Selection Criteria | 4, 7 | • Comparison of clinical characteristics and utilization when altering cohort definitions to assess potential bias, such as proportion of patients on hydroxyurea when requiring 1 vs 2 SS/S$\beta^0$ diagnoses |
| Expected Variables Present | 5, 8 | • Missingness in hydroxyurea prescriptions that triggered investigation for how to recover this information (using Source to Concept Vocabularies)<br>• Missingness in drug metadata across all institutions for hydroxyurea prescriptions |

## RESULTS

The results comprise our transparent reporting of data quality adjustments for this study as well as the downstream clinical results and findings.

### Data Quality Results

The data quality process described in Methods led to adjustments in the study design that we made prior to conducting our analyses, in order to more closely replicate the clinical intent of the original study. We describe these adjustments in Table 2, organized by summary of changes and rationale.

**Table 2.**

| Summary | Rationale |
| --- | --- |
| Final Cohort Definition:<br>1) $\geq 2$ diagnoses for SS/S$\beta^0$ at least 30 days apart;<br>2) $< 18$ at first SS/S$\beta^0$ diagnosis;<br>3) Evidence of hemoglobin electrophoresis or beta gene sequencing lab performed;<br>4) $\geq 3$ visits with a hematologist;<br>5) $\geq 2$ years of follow up; | • We recovered evidence of SCD phenotype tests but results were either missing or uninterpretable for several sites. Therefore, we considered evidence of test occurrence as one of many requirements for cohort inclusion.<br>• We modified criteria to include multiple SS/S$\beta^0$ diagnoses as a proxy for test-confirmed results.<br>• We required at least 3 visits with a hematologist and 2 years of follow-up to indicate ongoing care at an institution |
| Hydroxyurea drug utilization mapping greatly improved from AS-IS to DFE cohort | • We were able to recover hydroxyurea prescriptions by improving mapping between source values and standardized codes<br>• We also used DQ modules to improve face validity of proportion of patients taking hydroxyurea after adjustments to the cohort definition. |
| Duration of hydroxyurea use was modified due to missing prescription metadata | • We observed heterogeneity in days supply and daily frequency of hydroxyurea. As a result, we opted to use the better-populated drug exposure start and end dates across institutions. Imputation was only needed rarely. |
| Hematology specialty was improved from AS-IS to DFE cohort | • We were able to improve hematology specialty visits by expanding search terms and recovering unmapped specialty visits at some institutions |
| Lab results were more standardized and interpretable after DQ analysis | • We examined heterogeneity in lab units and results, and better standardized for analysis |
| Improved face validity of proportion of patients on hydroxyurea | • Initially our more sensitive cohort definition reduced the proportion of patients on hydroxyurea, particularly at one institution<br>• We required multiple SS/S$\beta^0$ diagnoses and ongoing interaction with a hematologist to reduce heterogeneity across institutions of patients eligible for hydroxyurea. |
| Hemoglobin testing and hydroxyurea prescriptions were observed across any visit, and not restricted to hematology clinic visits | • Specialty data was recoverable but many patients were seeking care outside of hematology, even after improving mapping for hematology clinic visits |

### Cohort Definition Changes

We began by using the original cohort definition to investigate attrition and utilization in PEDSnet (Figure 2). A variety of issues were discovered, ranging from gaps in hemoglobin typing result and medication prescription capture to inconsistent trends in hematology visits. For example, the original cohort demonstrated a sharp drop in eligible patients at the attrition step requiring a laboratory-confirmed diagnosis, with 2/10 sources dropping from the cohort entirely. In response, we reviewed test metadata and conducted key-word searches to augment standardized codes, allowing us to recover mismapped or mislabeled data. While this improved the discovery of SCD phenotype testing, results were still uninterpretable for some institutions. Because we were unable to recover these results or manually review patient records, we focused on diagnostic coding and utilization to determine appropriate proxy metrics. Requiring multiple diagnoses of SS/Sβ^0^ sharply improved face validity of utilization metrics and hydroxyurea prescriptions across institutions without excessively restricting the cohort and introducing potential bias. Figure 3 demonstrates the difference between the cohort definitions (1 vs 2+ SS/Sβ^0^ diagnosis events) across a series of utilization and demographic metrics, showing both overall impact of adapting study design to data characteristics as well as heterogeneity across institutions. Utilization increased near-uniformly across institutions but site E was most impacted. For example, site E inpatient and ED visits went from 0 and 0.83 visits per person year to 0.6 and 1.48 visits per person year, respectively. Similarly, study-related lab and drug availability also increased, with hydroxyurea prescriptions increasing from 18% to 64% of the cohort who received the drug. The final cohort definition is presented in Table 2.

**Figure 2.**
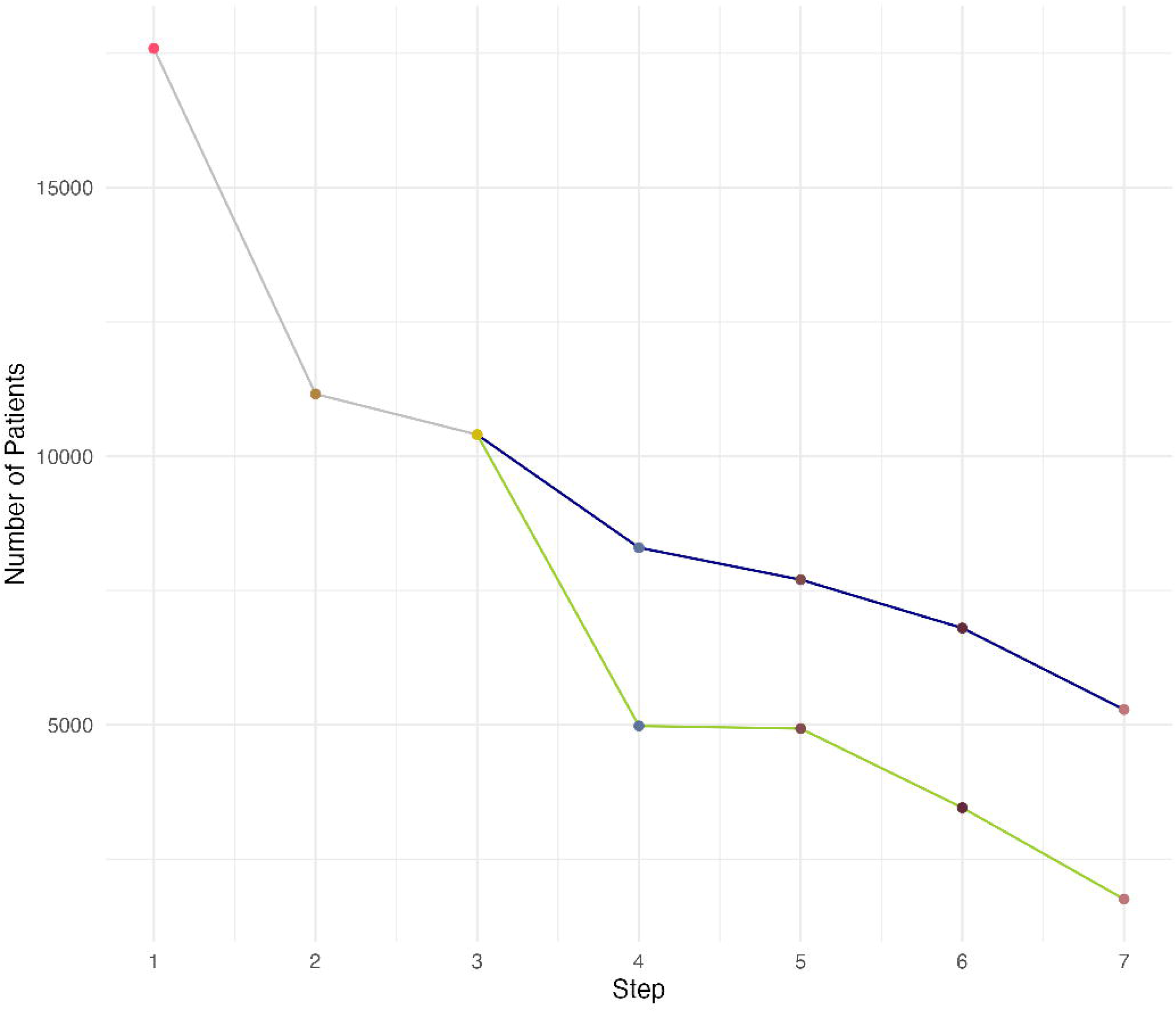
Effect of DQ on Cohort Attrition: Attrition steps common to both methods are shown in grey, those unique to the AS-IS arm in green, and those unique to the DFE arm in blue. <u>Common criteria</u>: 1) Patients with any SCD diagnosis code; 2) Patients with any SS/Sβ^0^ diagnosis code; 3) Patients with SCD-SS/ Sβ^0^ diagnosis code at <18 years old. <u>DFE-specific criteria</u>: 4) Patients with any SCD-related laboratory test result, after test remapping; 5) Patients with ≥3 outpatient visits with a hematologist, after specialty remapping; 6) Patients with ≥2 years of followup; 7) Patients with ≥2 SCD-SS/ Sβ^0^ diagnosis codes separated by >30 days. <u>AS-IS-specific criteria</u>: 4) Patients with SCD-confirmatory laboratory test result; 5) Patients with ≥3 outpatient visits; 6) Patients with ≥1 outpatient visit with a hematologist; 7) Patients with > 50% of outpatient visits with a hematology specialist.

**Figure 3.**
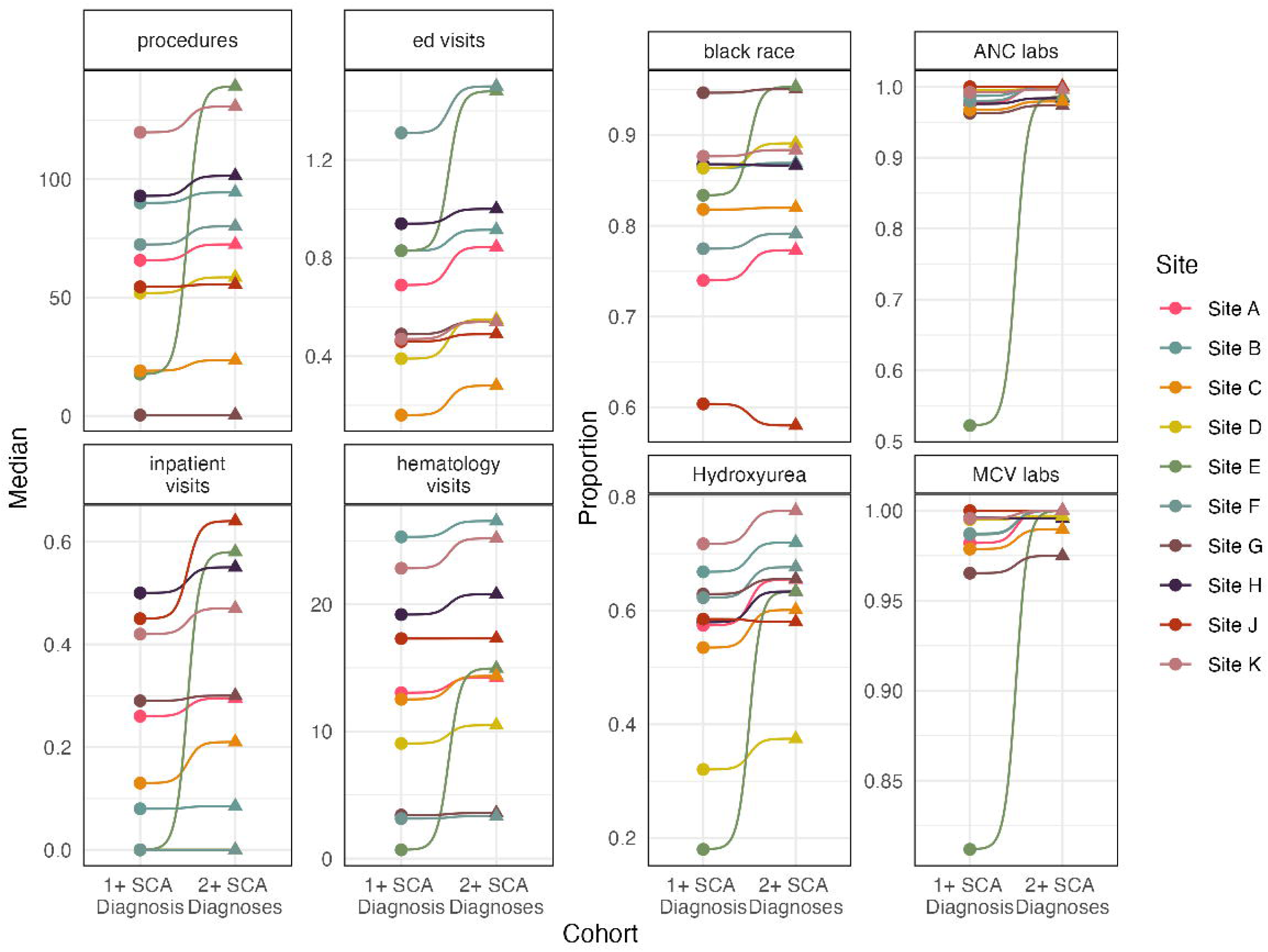
Sensitivity to Selection Criteria: Diagrams show effect of imposing a second diagnosis requirement on characteristics of the resulting cohorts, stratified by institution. Values for broad utilization measures are shown as median count per person-year for the cohort, while those for more study-related metrics are shown as proportion of cohort having at least one qualifying event.

### Additional Study and Variable Modifications

We focused on hydroxyurea utilization across our DQ process through two primary mechanisms. First, we reviewed the proportion of patients at different institutions who were prescribed hydroxyurea. Using SQUBA, we were able to longitudinally compare hydroxyurea prescription use across multiple cohort definitions. We initially observed high heterogeneity of hydroxyurea prescriptions; modifications to the upstream eligibility criteria produced more consistent hydroxyurea utilization across both time and institutions, by insuring a more consistent eligible population (Figure 3). Second, we observed that codes for hydroxyurea needed to be expanded, and improved mappings from source systems to recover missing hydroxyurea prescriptions.

When interrogating provider specialty information, we discovered anomalously high rates of other/unknown specialties and unexpectedly low rates of hematology specialty visits. To recover unmapped specialties and improve the rate of hematology specialty visits, we conducted a string search of unmapped values from institutional source systems (*source values* in the OMOP common data model) to identify missed mappings and were able to augment the standardized concepts. We were able to recover 64% of specialty encounters for 3 institutions with lowest mapping rates in our cohort. While the authors of the original study never stated the requirement for exposure or outcome definitions to be observed in hematology clinic, their analysis focused on utilization within hematology. Our analyses revealed that patients received care in a variety of settings, and therefore restricting observation to only hematology clinic visits would potentially bias our sample.

In investigating laboratory results data, we discovered that ANC values were most often reported in units of thousands per microliter. This unexpected unit prompted conversion to a standard, clinically acceptable unit for this result and others. We were also able to improve result interpretability across a range of tests including hemoglobin.

We discovered gaps in drug metadata such as days supply, daily frequency, refill amount, and quantity prescribed, which are required for computing duration of medication use. Figure 4 shows the heterogeneity of these metadata across institutions. Institutions may reliably capture one piece of metadata but not another to consistently and reliably compute duration of medication. For example, site J has nearly complete capture for refills (100%) and frequency (89%), but lower capture for quantity (57%) and days supply (0%). This is in contrast with Site H, which generally performed better on quantity and days supply rather than refills and frequency. Instead of imputing missing drug metadata, we opted to standardize more simply on drug exposure start and end dates associated with every drug prescription, which needed imputation much less often than other metadata because it was more consistently populated. In contrast, the original study was able to confirm duration based on manual chart review, which is not feasible in large multi-institutional network studies.

**Figure 4.**
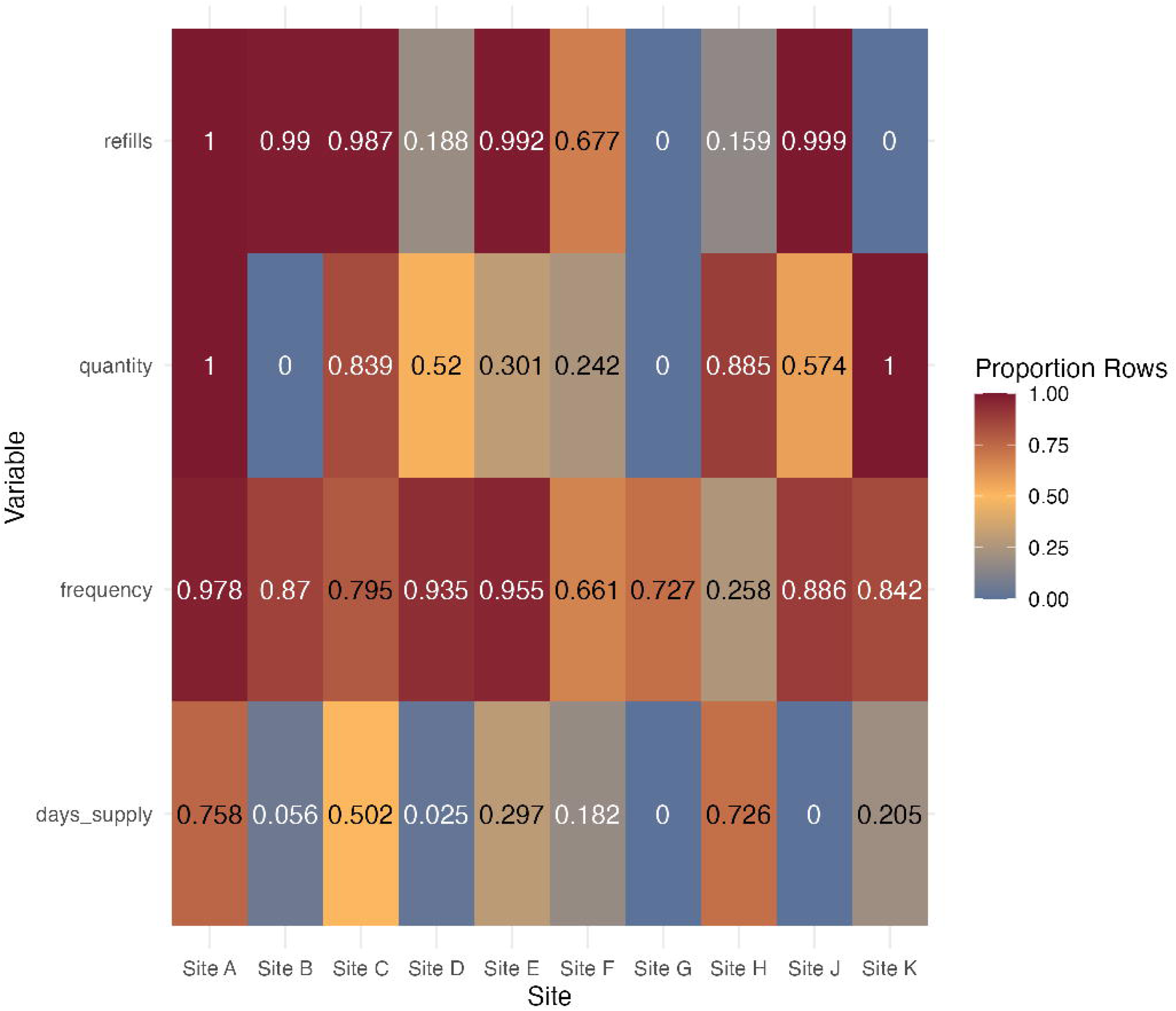
Variation in Drug Metadata by Institution: Heat map showing proportion of drug prescription records that include specific types of metadata used to infer duration of exposure.

### Clinical Results

The following descriptive statistics are reported in order of DFE, AS-IS, and CHOA. The total number of patients in each cohort are 5265, 1754, and 2147, respectively. The mean (median) age at cohort entry was 5.03 (2.83), 4.37 (1.76), and unreported for the CHOA cohort, while the length of follow-up in years was 5.79 (5.5), 5.30 (4.76), and 5.5.

**Table 3:**
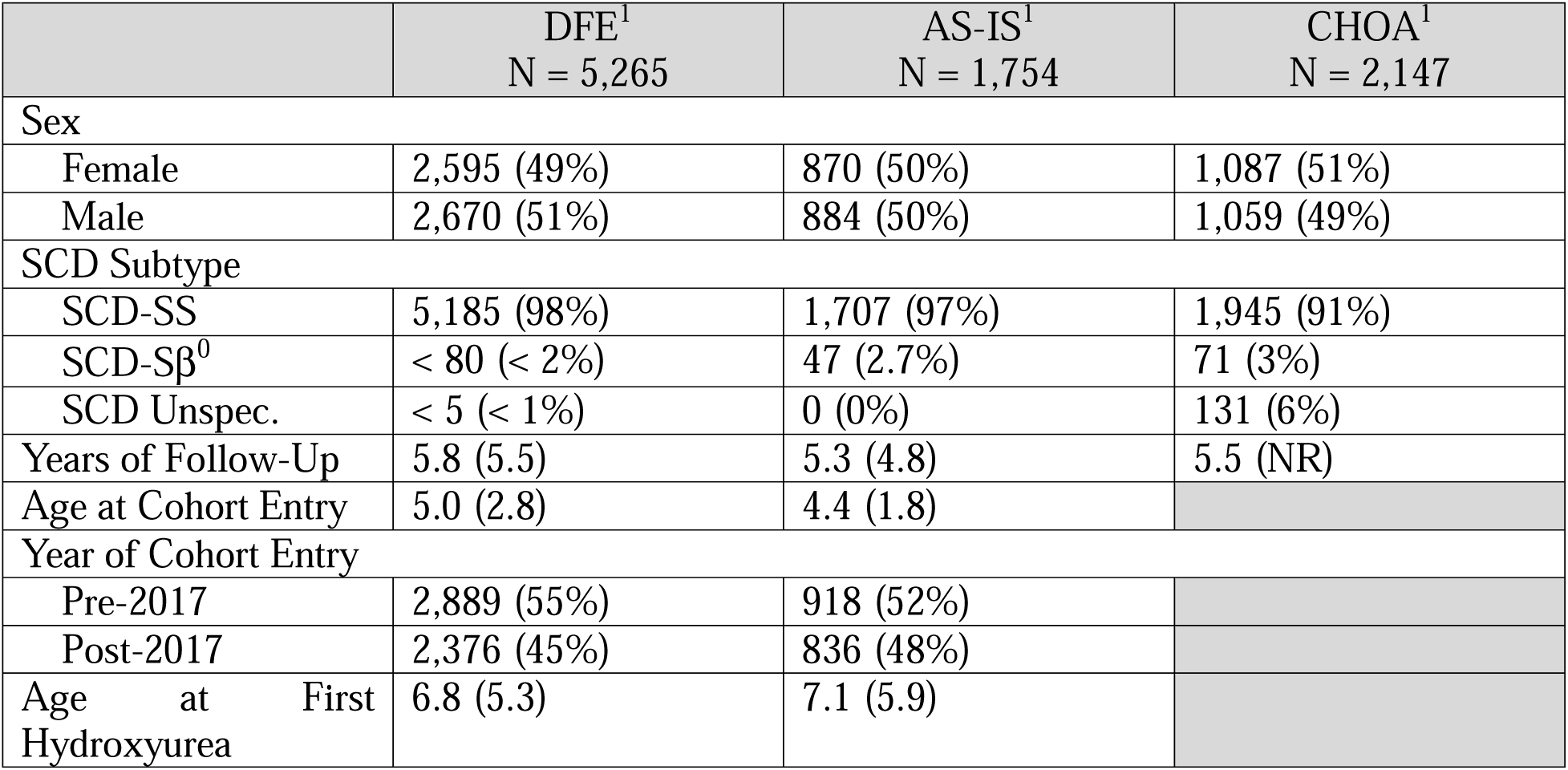
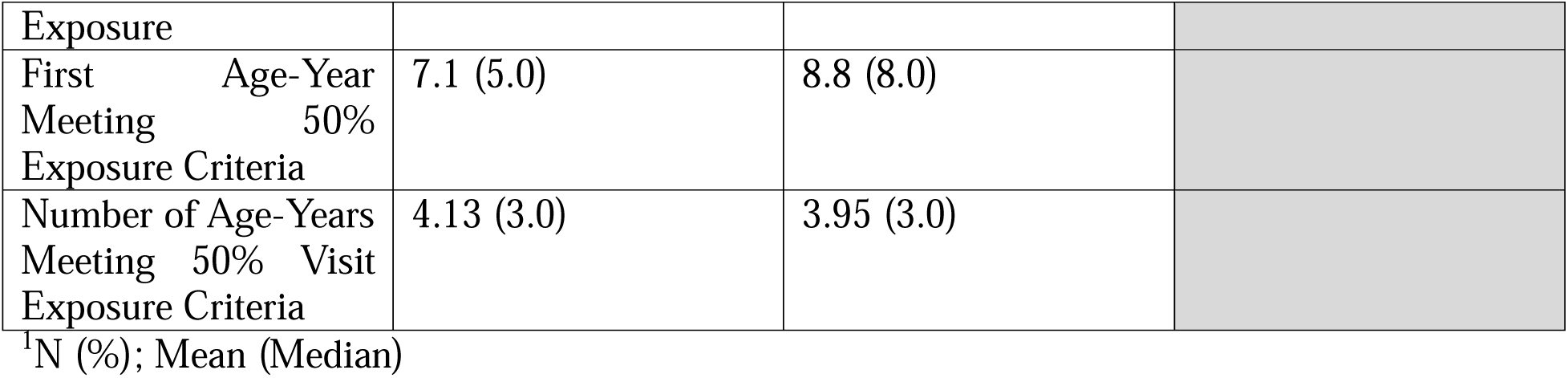
Cohort Descriptives.

We computed the same utilization metrics as the original study, including change in ED (Figure 5) and hospitalization days per age-year. The average treatment effect on the treated (ATT) in ED days per age-year for the AS-IS, DFE, and CHOA cohorts measured in change of visits per year were -0.08 (CI -0.26,0.09), -.44(CI -0.60,-0.26), and -0.36 (CI -0.57,-0.16), respectively, and the ATT for change in hospitalization days per year was -0.61 (CI -1.24, 0.02), -1.93 (CI -2.46, - 1.40), and -0.84 (CI -1.51, -0.17). We observe the same directionality and general magnitude in changes in our DFE and CHOA cohorts, but consistent effect attenuation in the AS-IS cohort for both measures.

**Figure 5.**
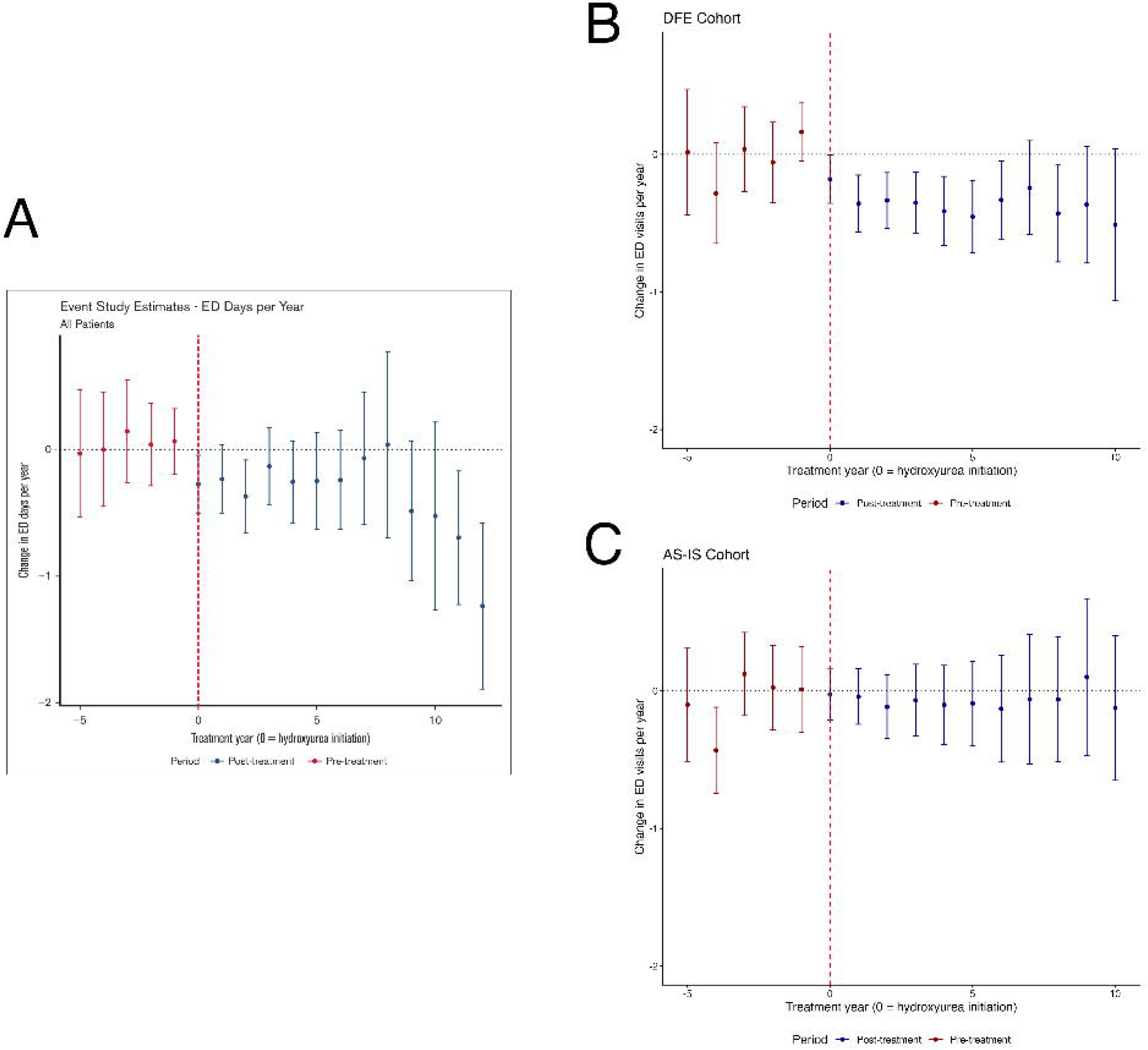
Difference-in-difference Analysis for ED Visits Per Year: (A) CHOA study (reproduced by permission from ^13^) (B) DFE arm (C) AS-IS arm

Finally, we computed change in average hemoglobin per age-year, for the full cohort as well as an “adherent” cohort, which included patients who met the following criteria: never treated with hydroxyurea (controls) or (a) treated for only one age year; (b) exhibited an average annual MCV value that was 110% of the baseline MCV value (recorded on the same date or within the year prior to hydroxyurea initiation). The authors of the original study used this cohort to attempt to exclude patients who did not demonstrate physiologic evidence of hydroxyurea use and were motivated to apply these criteria because they observed decreased effect for later treatment years. Figure 6 shows the hemoglobin labs for the full CHOA (Panel A) and DFE cohorts (Panel B) as well as the adherent cohorts. Both cohorts show protective trends of hydroxyurea on utilization, though the DFE cohort demonstrates continued protective effect for longer years of follow-up, particularly in the adherent cohort. The CHOA cohort showed greater increase overall [ATT 0.56 g/dL, (CI 0.39,0.73) vs [ATT 0.10 g/dL, (CI -0.03, 0.22)], though some of these differences may be attributable to age at initiation and dose titration, both of which were unavailable for comparison during analyses. The original study did not report on the ATT for the adherent cohort (though did demonstrate sustained benefit in follow up) but the effect on the DFE cohort was ATT .16 g/dL (CI -.03, 0.36), with early (year 1-2) and late treatment years (years 7-9) showing sustained improved hemoglobin over middle years In contrast, the AS-IS cohort demonstrated a negative impact of hydroxyurea in the overall cohort [ATT -0.44 (CI -0.67, -0.22)] and no effect in the adherent cohort [ATT -0.07 (CI -0.43. 0.30)].

**Figure 6.**
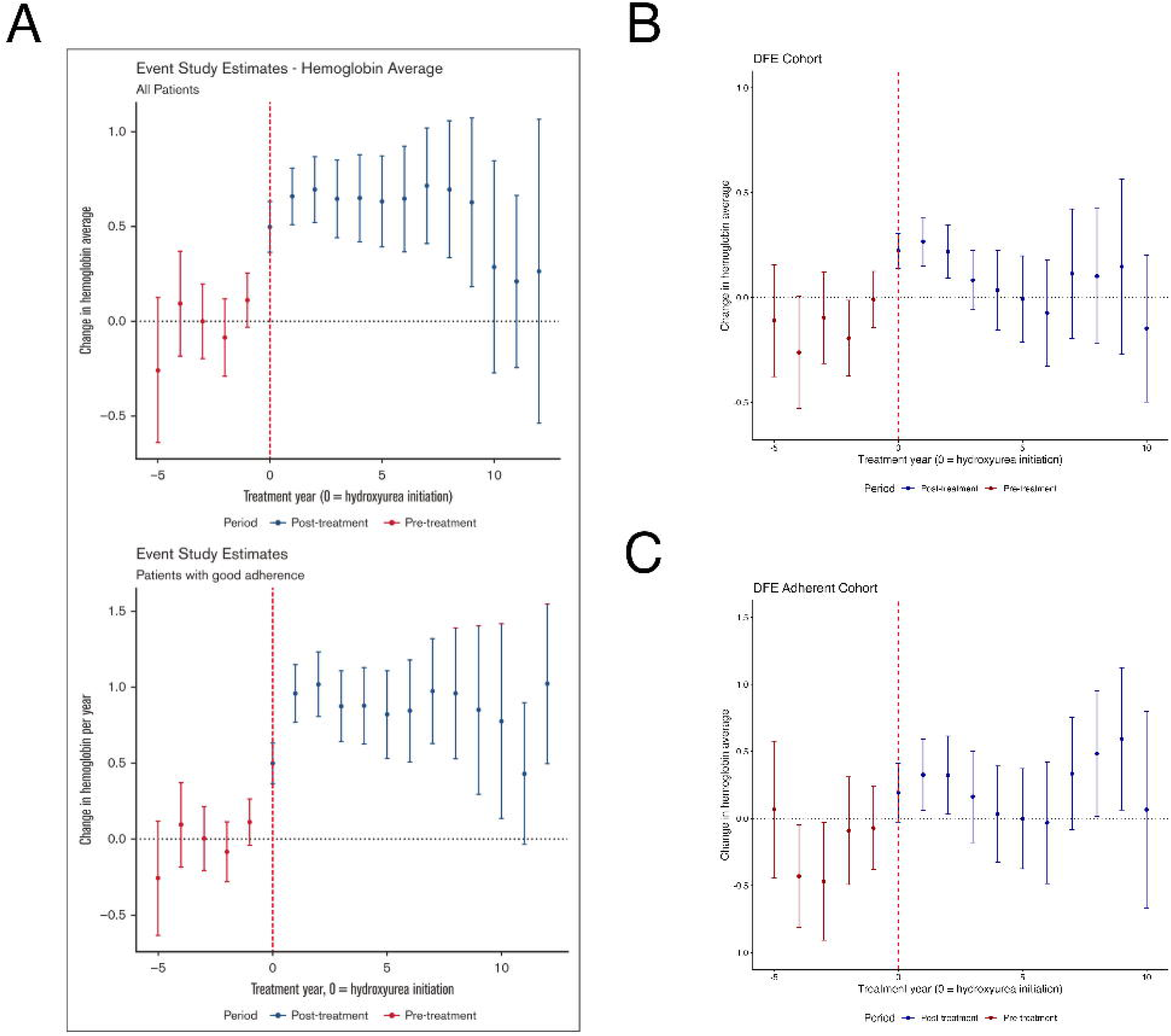
Difference-in-difference Analysis for Hemoglobin (g/dl): (A) CHOA study (reproduced by permission from ^13^) (B) DFE arm, all users (C) DFE arm, adherent users

The adherent DFE cohort also exhibited improved change in ED visits per year [ATT -0.55, (CI - 0.91, -0.19) and hospitalization-days per year [ATT -2.32 (CI -3.56, -1.08)] over the adherent AS-IS cohort, where change in ED visits per year and hospital-days per year were ATT 0.08 (CI - 0.20, 0.37) and ATT -.64 (CI -1.36, 0.07), respectively.

## DISCUSSION

This study demonstrates that replication of clinical research using real-world EHR data requires deliberate, study-specific data fitness assessment rather than routine translation of inclusion and exclusion criteria from study designs. Despite descriptive characteristics that were broadly plausible across the AS-IS, DFE, and CHOA cohorts in terms of demographics, follow-up duration, and age at entry, the analytic results diverged substantially. This finding underscores a critical and underappreciated principle: superficial comparability in cohort characteristics does not guarantee comparability in research-ready data quality. The same considerations will apply to target trial emulation and other methods that translate smaller, manually intensive study designs to large populations and datasets.

The use of standardized, modular DQ tools from the SQUBA package enabled systematic documentation of design decisions made in response to DQ findings, from specialty remapping to cohort definition refinements. This level of transparency is rarely achievable through *ad hoc* data curation and is largely absent from current reporting norms in clinical research. Journals require rigor in statistical reporting, but few standards exist for documenting how data were interrogated, curated, or modified prior to analysis. For replication studies in particular, this gap is consequential: without knowing what accommodations were made and why, results across studies cannot be compared.

The AS-IS arm illustrates what happens when large-scale data are used without this rigor. Even with minor modifications to accommodate the PEDSnet data environment, the AS-IS cohort produced consistently attenuated effects, a detrimental hemoglobin signal, and trends that lacked face validity. This demonstrates that the analytic and operational complexity of multi-institutional EHR data demands proportional methodological investment. The growing accessibility of large clinical databases has made it technically easy to apply study criteria at scale, but it does not substitute for domain knowledge, iterative data interrogation, or transparency in reporting.

The DFE cohort produced results with substantially greater validity. The ATT for ED visits and hospitalization days were larger in the DFE than CHOA cohort, potentially reflecting the power afforded by a cohort more than twice the size of the original study, as well as the geographic and demographic diversity of patients across ten PEDSnet institutions. Where CHOA captures the experience of a single specialized SCD center, PEDSnet captures a more representative cross-section of how hydroxyurea performs across varied real-world care settings. The consistency and strength of the utilization signal across this heterogeneous population reinforces the robustness of hydroxyurea’s clinical benefit and demonstrates that multi-institutional EHR data, when properly curated, can generate effect estimates with greater generalizability than single-center studies.

For hemoglobin, the DFE cohort showed a consistent protective direction of effect, with the adherent cohort demonstrating more sustained change over time, mirroring patterns observed at CHOA. However, the effect size was attenuated and the confidence intervals were wider, reflecting greater heterogeneity across institutions. Several factors may contribute to this. Practice variation across the ten PEDSnet institutions in hydroxyurea prescribing, dose titration, and monitoring intensity likely dilutes the average hemoglobin effect observed relative to a single center. Dosing information and age at first hydroxyurea exposure, both of which can substantially influence hemoglobin response, were not available from the CHOA cohort for direct comparison. It is also possible that the attenuated hemoglobin effect reveals a genuine finding, reflecting greater variation when using larger, multi-institutional datasets. The evidence for Hemoglobin F (HbF) is more biologically plausible and robust than total hemoglobin, with one clinical trial demonstrating improvements in the former but not the latter in patients treated with hydroxyurea and phlebotomy vs chronic transfusion and chelation^19^. However, HbF was not available for analysis in the original study, and further research is warranted to explore the real-world impact of hydroxyurea on HbF^20, 21^.

Taken together, these findings support the broader adoption of standardized data fitness frameworks as a prerequisite for replication research using real-world clinical data. The DQ process described here is not a bespoke solution; the SQUBA modules are open-source, domain-agnostic, and designed for reuse. The research community now has unprecedented access to very large clinical databases, including Cosmos, TriNetX, and others, that offer scale and efficiency infeasible a decade ago. Such access, while valuable, should not come at the cost of scientific rigor. Larger databases amplify both signal and noise, and without systematic data fitness assessment, the latter can easily masquerade as the former. As the field moves toward greater reliance on multi-institutional EHR networks for evidence generation, the infrastructure for transparent, reproducible data assessment must advance alongside it.

This replication study demonstrates that standardized, transparent data fitness assessment is essential, not incidental, to generating valid and replicable evidence from multi-institutional EHR data. Applying the original study’s criteria without adjustment produced attenuated and often implausible effect estimates, while systematic data fitness assessment yielded results that were both more concordant with the original single-center findings and more robust across a larger, more diverse patient population. These findings show that superficial similarity in cohort characteristics is not sufficient evidence of comparable data quality, and that data fitness assessment must be a required, reported component of study methodology rather than an *ad hoc*, undocumented step. Because the SQUBA modules used here are open-source and common-data-model agnostic, this approach generalizes beyond sickle cell disease and hydroxyurea to other conditions and multi-institutional research networks. Broader adoption of standardized data fitness reporting could meaningfully improve the reproducibility and credibility of real-world evidence.

## CRediT Author Contributions

**Hanieh Razzaghi**: Conceptualization, Methodology, Validation, Writing-Original Draft, Formal Analysis, Supervision **Kaleigh Wieand**: Formal analysis, Data Curation, Visualization, Writing-Original Draft **Annabel Pinkney**: Data Curation, Writing-Original Draft **Charles Bailey**: Methodology, Validation, Writing-Original Draft, Funding Acquisition, Supervision

## Acknowledgements

The research reported in this publication was conducted using data from PEDSnet, A Pediatric Clinical Research Network. PEDSnet has been developed with funding from the Patient-Centered Outcomes Research Institute (PCORI); PEDSnet’s participation in PCORnet is funded through PCORI award RI-CHOP-01-PS10. Support to develop the methods for this project was supported through PCORI Contract ME-2020C3-21199. The authors are grateful for the sharing of information by the patients, families, and institutions in PEDSnet, which makes more effective research possible.

## Data Availability

The clinical data underlying the results in this manuscript contain protected health information about individual patients, and therefore cannot be made publicly available. Reasonable requests for use of these data would require appropriate oversight by a research ethics board and use of data sharing agreements ensuring regulatory compliance and privacy protection. Requests can be made via https://pedsnet.org/work-with-us/collaboration-request/. The software used to create the visualizations can be found on GitHub at https://github.com/ssdqa/ssdqa_comp_paper3.

## Declaration of generative AI and AI-assisted technologies in the manuscript preparation process

During the preparation of this work, the author(s) used Claude for assistance with editing and flow of the manuscript. The authors used Claude to help streamline language, and they provided very specific directions and context. The author(s) reviewed and edited the output as needed and take full responsibility for the content of the published article.

